# Effect of Redcurrant (Ribes rubrum) fruit powder on patients with COVID-19, A randomized double-blinded clinical trial

**DOI:** 10.1101/2024.12.15.24319053

**Authors:** Azadeh Ahmadi Pirmorad, Sakineh Ebadipour, Amir Amiri-Sadeghan, Habib Zarredar, Milad Asadi, Majid khalili, Akbar Sharifi

## Abstract

**Introduction:** COVID-19 has affected many people worldwide. Effective treatments have been reported using medicinal plants which their multi functionality may beneficial. Considering the importance of innate immune response and pathologic effect of cytokine storm in COVID-19 patients, this study examined the effect of Redcurrant (*Ribes rubrum*) fruit powder capsules on inflammatory cytokines of IL-6 and TNFα as primary objectives. Moreover, its effect on the COVID-19 patients’ recovery symptoms was evaluated.

**Methods:** We analyzed the clinical effect on recovery symptoms, hematological parameters, and the serum levels of interlucin-6 and TNFα in the placebo-controlled and treated patients with Redcurrant fruit powder.

**Results:** Redcurrant fruit powder capsules significantly decreased pro-inflammatory cytokines of IL-6 and TNF-α, and reduced the clinical recovery time of COVID-19 and fatigue (from 7.3 ± 2.09 to 2.9 ± 1.28 days) as well as cough (from 5.67 ± 2.4 to 2.4 ± 1.7) days compared to the control group.

**Conclusion:** Redcurrant fruit powder might be a promising herbal treatment of COVID-19, and since no considerable side effect is observed in short-term usage, it might be beneficial to start treatment as early as possible in suspected cases of COVID-19.

## 1. Introduction

The pandemic of corona-virus disease 2019 (COVID-19) has affected more than 400 million people with approximately 5.8 million deaths worldwide (Organization, 2020). COVID-19 belongs to the distinct family of viruses which caused both Middle East respiratory syndrome and severe acute respiratory syndrome in the past (Wu et al., 2020). Since the outbreak of COVID-19, clinical trials of herbal medicine (Ang et al., 2020a) have been conducted alongside the modern medicine interventions, and among them, significantly effective treatments are documented (Ang et al., 2020b; Mirzaie et al., 2020; Yu et al., 2022). Moreover, in the most cases, an herbal formulation is not just a purified substance, and usually includes a whole part of a plant that may provide many bioactive phytochemicals each with different functionality that may interfere with the necessary elements of the virus life cycle (Alam et al., 2021). The multi-functionality of these formulations may bring some advantages in combating such pathogens.

Berries of different genus and species are considered as potential anti-influenza phytotherapy in both prevention and treatment. In addition to providing health-essential substances like vitamin C and or carotenoids, the bioactive compounds including polyphenols (anthocyanins, flavonoids, proanthocyanidins and acids) and polysaccharides exert some effects such as suppressing replication system of the virus both either directly or indirectly, e.g. through blocking surface glycoproteins of the virus together with modulating the immune system (Gramza-Michałowska et al., 2017). It is shown that the anti-inflammatory effects of herbal polyphenols is beyond their antioxidant effect, and direct activation or inhibition of various cell signaling processes such as the activation of transcription factors, altering receptor activation as well as possible direct ligand activity are also involved in this process (Scalbert et al., 2005).

The health-promoting effects of various formulations including berries are shown in many clinical trials. For instance, Sambucol is a preparation containing elderberry juice, raspberry extract and honey; when administered for 6 days, it reduced ailments caused by infection with influenza type A and type B viruses in patients had been suffering from at least 3 influenza symptoms for 24 h. Significant improvement in health started as early as 3–4 days after beginning the treatment which was prolonged about two times in the placebo group (Zakay-Rones et al., 2004). In another study, pastilles of elderberry extract reduced the severity of systemic symptoms and nasal symptoms within 24 h of the onset of treatment (Kong, 2009).

Moreover, direct interaction of the anthocyanins with the virus is reported. For example, molecular docking studies have shown that anthocyanin derivatives occupy the pocket area of active site of main protease of Covid-19, which is an essential enzyme in the virus life-cycle (Abdjan et al., 2020; Akinnusi et al., 2022). The study showed that cyanidin-3-rutinoside can be an appropriate candidate as an anti-COVID-19 drug.

Iranian Qara Qat fruit (redcurrant, *Ribes rubrum*) in Arasbaran forests, traditionally, is considered as an herbal drug to prevent and treat common cold and influenza by the native people (Mollaamin et al., 2021). The anthocyanin content of various berries was determined in a study that confirms the presence of cyanidin-3-rutinoside in redcurrant fruits (Jara-Palacios et al., 2019). Therefore, the consumption of this fruit is probable to interrupt the viral replication and harness its activity. Besides, the immunomodulatory effects of anthocyanins may be in accordance with the healing process. Considering the positive outputs and the statements from the individuals who consumed this fruit when they were infected by COVID-19, we decided to investigate the effect of the fruit of *Ribes rubrum* in a placebo-controlled randomized trial on COVID-19 patients.

## 2. Methods

### 2.1. Study design

This study is a randomized clinical trial in adherence to CONSORT guidelines, and is a parallel group designed to evaluate the effectiveness of the fruits of redcurrant on COVID-19 patients.

### 2.2. Ethical considerations

The study process was explained and the written informed consents were admitted and signed by the volunteer patients, before entering the study. Throughout the study, the researchers adhered to the principles of the declaration of Helsinki and confidentiality of patient information. All costs of the project were borne by the researchers and patients did not incur any costs.

### 2.3. Sample size, Inclusion and Exclusion criteria

Assuming confidence limits of 95%, power of 80%, equal number of samples in each group, and probability of exposure in control and case group as 0.1 and 0.4 respectively, and drop-out rate of 30%, 80 subjects has been identified. Simple randomization was applied using ID card number of patients to be odd or even. Trial was blinded for both patients and researcher, and bottles named as ‘A’ and ‘B’ were administered for odd or even ID card numbers respectively.

Inclusion criteria:

Real-time PCR diagnosed outpatients with stable vital signs, and blood oxygen saturation higher than 95% without any other underlying diseases such as diabetes, cardiovascular, blood pressure, kidney failure, liver disease could enter the study. Obeying this criterion, no patient with fast breathing or difficulty in breathing was entered the study. Obviously, patients with any severe symptoms such as severe respiratory distress syndrome, and patients admitted to intensive care units were not included in the study. All patients were aged 18 or older, and not pregnant or breastfeeding.

Exclusion Criteria: patients feeling dissatisfaction with study participation, dissatisfaction with continuing herbal supplements for any reason, and patients with incomplete questionary were not involved in endpoint measurements.

### 2.4. Herbal capsules and dosage

Capsules containing one gram of *Ribes rubrum* powder were provided by knowledge-bases company active in medicinal plant production. The plant taxon was confirmed by expert botanist and also by sequence of 16s rDNA of chloroplast, which was in accordance with accession number of NC_080516.1 in complete genomic molecules of NCBI, and previously reported classifications (Senters and Soltis, 2003; Sun et al., 2023). The daily intake of powder dose as (6 g/day) was extracted from previous studies on other berries (Gramza-Michałowska et al., 2017).

### 2.5. Measured parameters

A scaled questionnaire covering the levels of fever, coughing, body aches, diarrhea, anorexia, sense of smell and taste, and generalized weakness were filled by the patients through a daily flowing up. The blood and serum tests including White Blood Cell (WBC), Red Blood Cell (RBC), Hemoglobin (Hb), Platelets (Plt), C-reactive Protein (CRP), Interleukin (IL-6), Tumor Necrosis Factor (TNFα), Erythrocyte Sedimentation Rate (ESR), and Lactate Dehydrogenase (LDH) were carried out at the first day and on day fourteen. The treatment group received a total dose of 6 g/d of encapsulated redcurrant fruit powder (three times a day, two 1g-capsules each time), and the placebo block received starch in the same shaped and color capsules.

### 2.6. Statistical analysis

Descriptive and inferential statistics including the Fisher’s exact test, Chi-square and independent T-tests, were performed to analyze the demographic and clinical data. The laboratorial and clinical data were compared parametrically using repeated measures analysis and mixed design analysis of variance (ANOVA). SPSS 16.0, and GraphPad Prism were used for data analysis and designing the graphs. P-values lower than 0.05 were considered as significant difference.

## 3. Results and discussion

The blood test included the measurements reported to be predictive or valuable in Covid-19 patients. Several hematological parameters have been shown to be associated with COVID-19 infection and severity such as platelets, white blood cell total count, lymphocytes, neutrophils, and hemoglobin. Decreased platelet, lymphocyte, hemoglobin, eosinophil, and basophil count, increased neutrophil count and neutrophil-lymphocyte and platelet-lymphocyte ratio have been associated with COVID-19 infection and worse clinical outcomes (Palladino, 2021). Moreover, higher levels of CRP indicated more severe infection and have been used as an indicator of COVID-19 disease severity (Chen, W. et al., 2020; Stringer et al., 2021) Also, Lactate Dehydrogenase (LDH) level is shown to be an independent risk factor for the severity and mortality of COVID-19 (Li et al., 2020). Due to the mentioned merits, these factors were measured on the first (day1) and fourteenth day (day14) after the patient entered the study as summarized in Table1. Treatment with redcurrant powder with the dose of 6 g/d did not significantly (p<0.05) affect the measured biomarkers. However, considering the p-values < 0.1, a decreasing effect on LDH and WBC were observed. Therefore, the potential of the treatment to reduce the tissue damage, and harnessing the infection is probable, and further trials with higher number of entered patients will elucidate this effect more confidently. Besides, the scores of fatigues and coughs have dropped to their half initial values faster in treatment group than control. fatigue score dropped in 7.3 ± 2.09 days in control group to 2.9 ± 1.28 days in treat group and the scores of coughs dropped in 5.67 ± 2.4 days in control group to 2.4 ± 1.7 days compared to the treat group. This effect was clinically meaningful, in spite of the fact that measured quantities of Table1 were not significantly different in control and treated patients.

**Table 1:** Blood test results of the patients in control and treatment groups on entering day (day1) and 14 days after (day14)

| Test | Placebo Day1<br>Mean (SD) | Placebo Day14<br>Mean (SD) | Treatment<br>Day1<br>Mean (SD) | Treatment<br>Day14<br>Mean (SD) | P-value<br>Placebo-Treatment<br>Day14 |
| --- | --- | --- | --- | --- | --- |
| WBC ($\times 10^3/\text{ml}$) | 10750 (5186) | 8582 (4079) | 10712 (5053) | 8892 (3097) | 0.08 |
| Lymph (%) | 10.33 (6.64) | 10.60 (8.25) | 9.60 (6.75) | 9.94 (7.34) | 0.67 |
| Plt$\times 1000$ | 360 (46.41) | 211.27 (72.88) | 213.75 (76.19) | 207.88 (92.24) | 0.34 |
| CRP (mg/L) | 81.4 (66.5) | 54.93 (47.7) | 93.4 (66.2) | 53.02 (43.0) | 0.88 |
| Hb | 12.86 (1.91) | 13.07 (1.9) | 12.63 (2.77) | 12.13 (3.75) | 0.75 |
| ESR | 54.54 (30.11) | 37.92 (30.65) | 48.23 (29.9) | 35.62 (21.85) | 0.79 |
| LDH | 1055.4 (434) | 913.8 (409.24) | 831.81 (391.9) | 685.77 (391.25) | 0.06 |

Cytokine storm (CS) is mentioned as the major hallmark of the COVID-19 disease, which is an aggressive hyperinflammatory immune response (Moore and June, 2020). CS is characterized by elevated serum levels of proinflammatory cytokines and chemokines, namely, IL-1, IL-6, IL-12, IFN-c and TNF-α (Du et al., 2021; Shimizu, 2019). A study of a longitudinal serum cytokine analysis of 207 COVID-19 patients showed that in very early inflammatory responses, IL-6, TNF-a, IL-10, and IL-1b rose in those with more severe disease (Bergamaschi et al., 2021). IL-6 has a pivotal role and is a double-edged sword in viral infection. IL-6 homeostasis governs the outcomes of immune-protection versus immunopathology. Therapeutic approaches that would attenuate the pro-inflammatory response without affecting the anti-inflammatory signaling process of IL-6 are preferred. ADAM10, a metalloproteinase switches the anti-inflammatory pathway of IL-6 to proinflammatory pathway by converting the membrane-bound form of IL-6 receptor to soluble type of IL-6 receptor, and plays a key role in developing the CS. However, raised IL-6 levels were associated with acute respiratory distress syndrome, increased requirement of mechanical ventilation, prolonged hospital stay, worse sequential organ failure assessment score, multiple organ impairment and intensive care unit admission (Chen, X. et al., 2020; Shekhawat et al., 2021; Zhou et al., 2020). As shown in Fig.1, IL-6 levels were significantly lower in treated patients respect to the placebo group. We did not elucidate how the treatment reduced the IL-6 level, which would help to understand if it may be a candidate supplement for preventing too or it just works after the disease is developed. However, the potential of the treatment in reducing the IL-6 level as a key cytokine in developing CS is demonstrated.

**Fig. 1.**
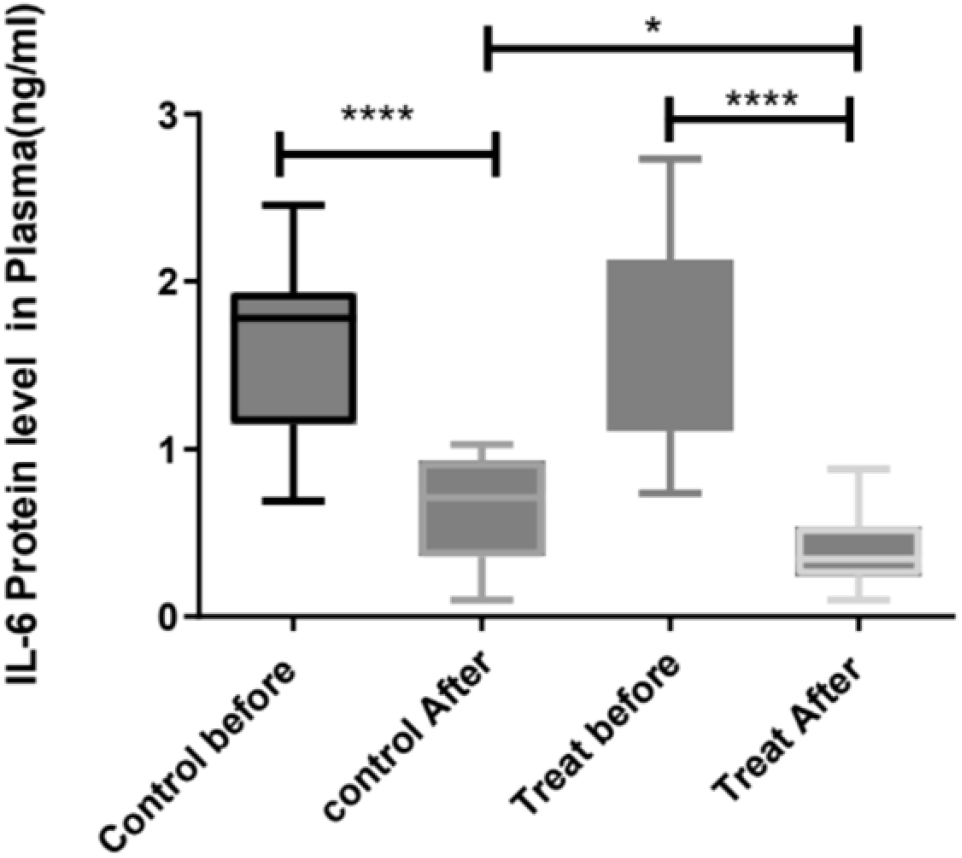

The comparison of plasma levels of IL-6 on day1 and day14 in placebo and treat-received patients. Shows that the treatment is significantly effective in the reduction of IL-6 levels in plasma

TNF-α is a major pleiotropic mediator of acute and chronic systemic inflammatory responses. It can simultaneously regulate cells apoptosis and proliferation while promoting the production of other chemokines and cytokines. It is also involved in a series of physiological processes such as anti-tumor responses, control inflammation, and immune system homeostasis (Croft, 2009). TNF-α is also one of the most important pro-inflammatory cytokines of the innate immune response, dysregulated TNF-α signaling can trigger CS. Although TNF coordinates the inflammatory response during the acute phase of inflammation, excessive TNF will suppress the immune system with the development of disease, which may lead to the adverse result (Aggarwal, 2003). Anti TNF-α therapy of COVID-19 have shown to be successful by some clinical trials (Guo et al., 2022). Here, as shown in Fig.2 treating with redcurrant powder decreased the TNF-α, therefore, it is effective to harness the development of CS.

**Fig. 2.**
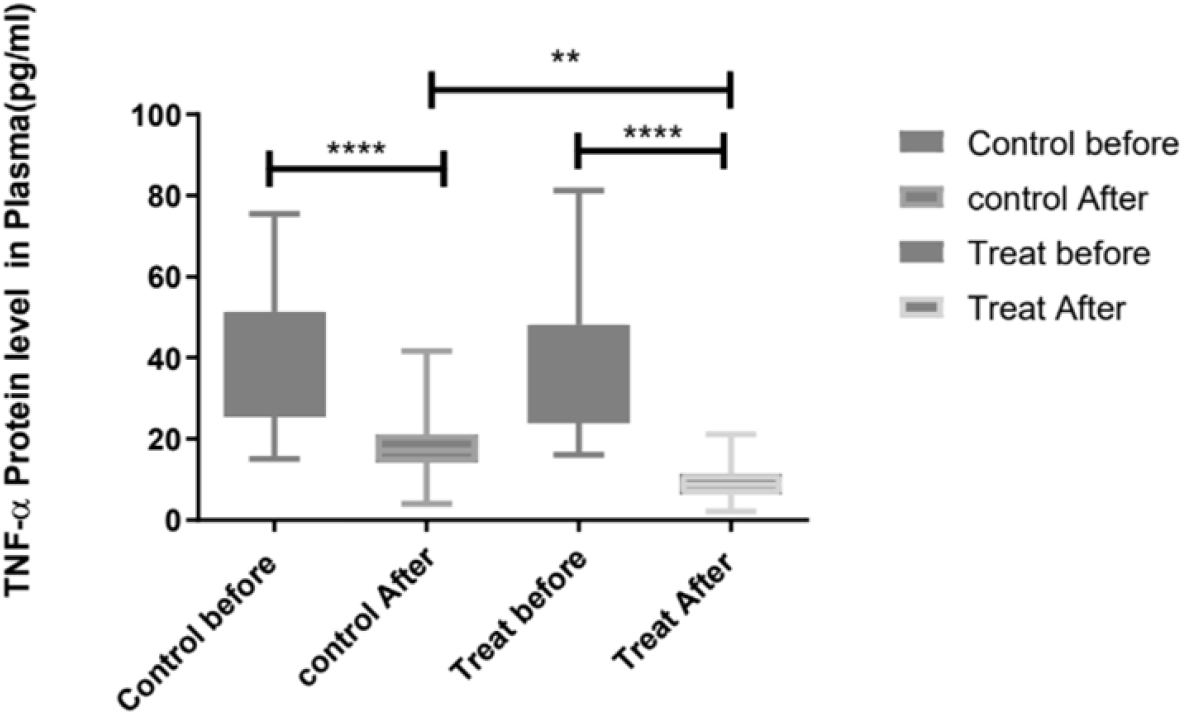

The comparison of plasma levels of TNFα on day1 and day14 in placebo and treat-received patients. Shows that the treatment is significantly effective in the reduction of TNFα levels in plasma

## 4. Conclusion

This study showed the effectiveness of using red currant powder in treating COVID-19 patients. Down regulating of IL-6 and TNF-α may be effective in harnessing the cytokine storm.

## Data Availability

All data produced in the present work are contained in the manuscript

